# Socioeconomic status, healthcare access, and risk of atrial fibrillation: an investigation using the Jackson Heart Study Research Materials

**DOI:** 10.64898/2026.09.09.26362592

**Authors:** Alvaro Alonso, Amit J. Shah, Chang Liu, Oluseye Ogunmoroti, Tené T. Lewis

## Abstract

**Background:** Rates of atrial fibrillation (AF) are lower in Black compared to White Americans. Socioeconomic disadvantage and reduced healthcare access may contribute to lower observed AF incidence in Black populations due to underdiagnosis and underascertainment. Therefore, we evaluated the association of socioeconomic status and healthcare access with AF incidence in a cohort of Black Americans.

**Methods:** 3,240 participants from the Jackson Heart Study, which recruited Black adults from the Jackson metropolitan area in 2000-2004, were followed up through 2016 for incident AF. Socioeconomic position (education and income) and healthcare access (insurance status, difficulty obtaining care, trust in providers, and satisfaction with care) were assessed at baseline. AF was ascertained from study electrocardiograms at baseline and exam 3 and self-reported physician diagnosis at annual follow-up surveys. Multivariable Cox regression was used to estimate hazard ratios (HR) and 95% confidence intervals (CI) for the association between exposure variables and AF incidence adjusting for potential confounders.

**Results:** During a mean follow-up of 11.5 years, 817 incident AF cases were identified. Higher educational attainment and income were associated with a reduced risk of AF. Compared with participants who had less than a high school education, those who attended trade or vocational school or college had an 18% lower estimated hazard of AF (HR 0.82, 95% CI 0.67, 1.02). Likewise, participants in the highest income category had a 39% lower estimated hazard of AF (HR 0.61, 95% CI 0.47, 0.78) compared with those in the lowest. Easier access to healthcare and greater satisfaction with received healthcare were also associated with lower AF risk.

**Conclusion:** Higher socioeconomic status and better healthcare access were associated with lower risk of AF in a cohort of Black Americans. These findings do not support a simple explanation in which socioeconomic disadvantage and poorer healthcare access lead to lower observed AF incidence through underascertainment within Black adults.

## INTRODUCTION

Atrial fibrillation (AF) is a common cardiac arrhythmia and an established risk factor for stroke, heart failure, dementia and overall mortality. Epidemiologic research over the last two decades has established the major risk factors for AF risk, including older age, obesity, hypertension, diabetes, and other cardiovascular diseases, such as heart failure or coronary artery disease.^1^ Prior work has described differences in the risk of developing AF across distinct racial and ethnic groups in the United States.^2^ In contrast to the patterns observed for most major cardiovascular risk factors and cardiovascular diseases, with more pronounced burden in Black than White populations,^3^ multiple studies have reported higher AF incidence in White than Black individuals.^2, 4^ This difference is paradoxical given the higher prevalence of most AF risk factors in the latter group. The factors underlying this paradox are unknown; however, several mechanisms have been advanced to explain it, including underascertainment of AF in Black individuals compared to other groups due to socioeconomic differences in income, education, healthcare access and healthcare utilization,^5^ factors correlated with genetic ancestry increasing risk in individuals of European ancestry,^6, 7^ or selective survival in Black individuals due to higher mortality rates from other cardiovascular diseases.

Prior studies have examined socioeconomic differences in AF risk in multiracial cohorts,^8, 9^ but socioeconomic gradients specifically within Black populations remain less well characterized. The Jackson Heart Study (JHS) provides a unique opportunity to evaluate multiple dimensions of socioeconomic status and healthcare access in a large Black cohort, without conflating them with the broader social and structural factors underlying Black–White differences in cardiovascular health. We therefore evaluated the associations of socioeconomic position, assessed by education and income, and healthcare access, assessed by insurance status and patient-reported measures of access and care, with incident AF in the JHS.

## METHODS

### Study population

The JHS is an ongoing community-based cohort to study the epidemiology of cardiovascular disease in Black Americans. The study recruited 5,306 adults 21-96 years of age from the Jackson, Mississippi, metropolitan area between 2000 and 2004. Additional in-person exams were conducted in 2005-2008 (exam 2) and 2009-2013 (exam 3). The aims and design of the JHS have been detailed elsewhere.^10^ For this analysis, we excluded participants who did not consent to share data beyond JHS investigators, those with missing electrocardiogram (ECG) or prevalent AF or atrial flutter at baseline, and those with missing information on education or income for a final sample size of 3,240 (**Figure 1**). The Jackson Heart Study was approved by the institutional review boards of Jackson State University, Tougaloo College, and the University of Mississippi Medical Center, Jackson, Mississippi. All study participants gave written informed consent. This analysis did not require review by the Emory University Institutional Review Board because the use of de-identified secondary data is not considered human subjects research. This manuscript was prepared using JHS Research Materials obtained from the National Heart, Lung, and Blood Institute (NHLBI) Biologic Specimen and Data Repository Information Coordinating Center (BioLINCC) and does not necessarily reflect the opinions or views of the JHS or the NHLBI.

**Figure 1.**
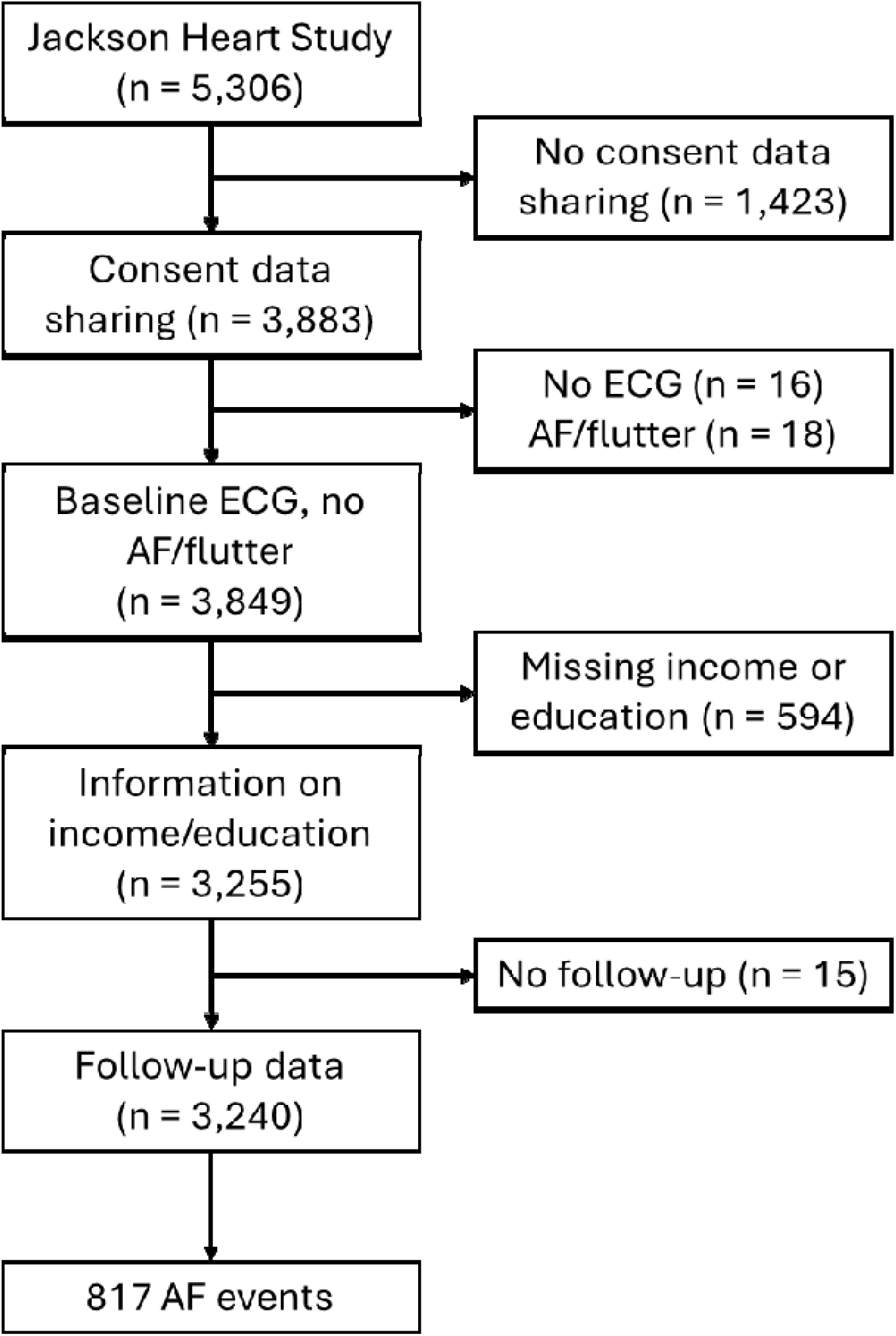
Flowchart of study participants, Jackson Heart Study, 2000-2016.

### Socioeconomic status and healthcare access assessment

Information on socioeconomic status and healthcare access was self-reported by study participants at the baseline examination using standardized questionnaires. Participants were asked about highest degree or years of school they had completed (including trade or vocational school, or college) or obtaining a GED if <12 years of school. Education was categorized as (1) less than high school, (2) high school graduate/GED, or (3) attended trade or vocational school, or college. Income was categorized by JHS as poor, lower-middle, upper-middle, or affluent based on self-reported annual family income, family size, and poverty threshold for the year of the examination. Insurance coverage was self-reported and categorized as uninsured, private insurance, public insurance (including Medicare, Medicaid, Veterans Administration), or dual public-private insurance. Participants were also asked about trust in medical care providers (“Thinking about the place you usually go for help with your medical problems, in general, how much do you trust them to take good care of you? Do you trust them very much, somewhat, not very much, or not at all?”), difficulty obtaining needed health services (“Overall, how hard has it been for you to get health services you have needed? Would you say it has been very hard, fairly hard, not too hard, or not hard at all?”), and satisfaction with their regular or most recent doctor or health professional (“Overall, how satisfied are you with your regular (or most recent) doctor or health professional? Would you say you are very satisfied, somewhat satisfied, somewhat dissatisfied, very dissatisfied, or not sure?”). Responses to these questions were dichotomized as “very much” or other response (trust), “not hard at all” or other response (difficulty), and “very satisfied” or other response (satisfaction).

### Ascertainment of atrial fibrillation

AF was ascertained from two sources: 12-lead ECGs done at baseline and exam 3, and self-reported physician diagnosis of AF at annual follow-up surveys. At baseline and exam 3, participants underwent a supine 10-second 12-lead ECG recording using a Marquette MAC/PC ECG recorder (Marquette Electronics). AF or atrial flutter were flagged using Minnesota codes. Participants (or a proxy / informant) were contacted annually via phone. As part of the annual follow-up, they were asked if they had received a medical diagnosis of AF (“Since we last contacted you [name], has a doctor said you [name] had an irregular heartbeat called atrial fibrillation, or atrial fibrillation on a heart scan or electrocardiogram tracing?”). An affirmative answer was considered as a diagnosis of AF, and the date of the annual follow-up was considered the diagnosis date. A secondary definition of self-reported AF to increase specificity required affirmative responses in at least two follow-up surveys, with time of incident AF defined as the second self-report. Follow-up surveys through 2016 were available for this analysis.

### Other covariates

Covariates were selected a priori based on prior literature and a conceptual framework identifying demographic characteristics, established AF risk factors, and social and psychosocial factors potentially associated with both socioeconomic status and AF risk. Age, sex, smoking status, and use of medications were self-reported at baseline. Height and weight were measured during the baseline exam, and body mass index was calculated as weight in kilograms divided by height in meters squared. Diabetes was defined as fasting serum glucose ≥126 mg/dL, use of glucose-lowering medications within two weeks of the study visit, or prior physician-diagnosed diabetes. Prior history of myocardial infarction was based on self-report or electrocardiographic evidence of a prior myocardial infarction (presence of major Q waves that met the specific standards or the combined presence of smaller Q waves and significant ST–T–wave abnormalities).^11^ Experiences of discrimination were assessed with the JHS discrimination instrument, which identifies experiences with and reactions to perceived everyday and lifetime discrimination.^12^ Everyday discrimination assessment was based on the scale by Williams et al.^13^ and ranged from 1 (no discrimination) to 7 (high discrimination). Evaluation of lifetime discrimination was adapted from the work of Krieger and Sidney,^14^ and Krieger,^15^ ranging from 0 (no discrimination) to 9 (high discrimination.) Discrimination was included as a relevant psychosocial factor because experiences of discrimination vary by socioeconomic position and have been associated with cardiovascular health through behavioral and physiologic stress-related pathways.^16^

### Statistical analysis

Baseline characteristics are reported by baseline education. Cox proportional hazards regression models evaluated associations between exposures of interest and AF incidence. Time to event was defined as the time from the baseline exam to AF incidence or censoring due to lost to follow-up, death, or latest available follow-up data. For each exposure, Model 1 adjusted for age and sex. Model 2 additionally adjusted for education, income, and insurance status, excluding the variable being evaluated as the primary exposure. Model 3 further adjusted for the remaining patient-reported healthcare access measures and measures of discrimination, again excluding any variable being evaluated as the primary exposure. Model 4 additionally adjusted for the main clinical risk factors for AF (height, body mass index, current smoking, systolic and diastolic blood pressure, use of antihypertensive medication, diabetes, and prior history of myocardial infarction.)^1^ Prior history of heart failure was not considered since that information was not collected at baseline. Model 2 was considered the primary confounder-adjusted model; Models 3 and 4 were used to evaluate the extent to which associations were attenuated after adjustment for potentially intermediate psychosocial, healthcare, and clinical factors. We conducted sensitivity analyses using a more specific definition of AF that required self-reported AF in at least two follow-up surveys. Finally, we conducted stratified analyses by sex (male, female) and age (<55, ≥55, approximate median age) to assess effect measure modification. Missing values in covariates (<5% for most variables) were imputed using the median value for continuous variables and mode for categorical variables. Cox proportional hazards assumption was tested using Schoenfeld residuals and plotting complementary log-log curves. No evidence of violations of the proportional hazards assumption for the main independent variables was observed.

## RESULTS

Of the 5,306 participants in JHS, 3,883 consented to share data with outside investigators. After excluding individuals with prevalent AF or atrial flutter at baseline (n = 18) and those without a baseline electrocardiogram (n = 16), 3,849 participants remained eligible. We further excluded participants with missing information on education or income (n = 594), yielding 3,255 individuals; an additional 15 participants without follow-up information were excluded, resulting in a final analytical sample of 3,240 (**Figure 1**).

Participants were followed up for up to 17.3 years after baseline. Over a mean (median) follow-up of 11.5 (13.6) years, we identified 817 incident AF events (of which 428 met the more stringent secondary definition requiring two self-reports during follow-up), corresponding to an incidence rate of 22 per 1,000 person-years. **Table 1** shows the participants’ characteristics by level of education. Participants with less than high school education and no GED were older, had a higher burden of cardiovascular risk and disease, and experienced greater socioeconomic disadvantage. They were more likely to have lower income, report more difficulty obtaining healthcare, and express lower satisfaction with their care. Participants in this group reported lower everyday and lifetime discrimination than those with higher education. Patterns were similar when evaluating participants’ characteristics by income level (**Supplemental Table S1**).

**Table 1.**
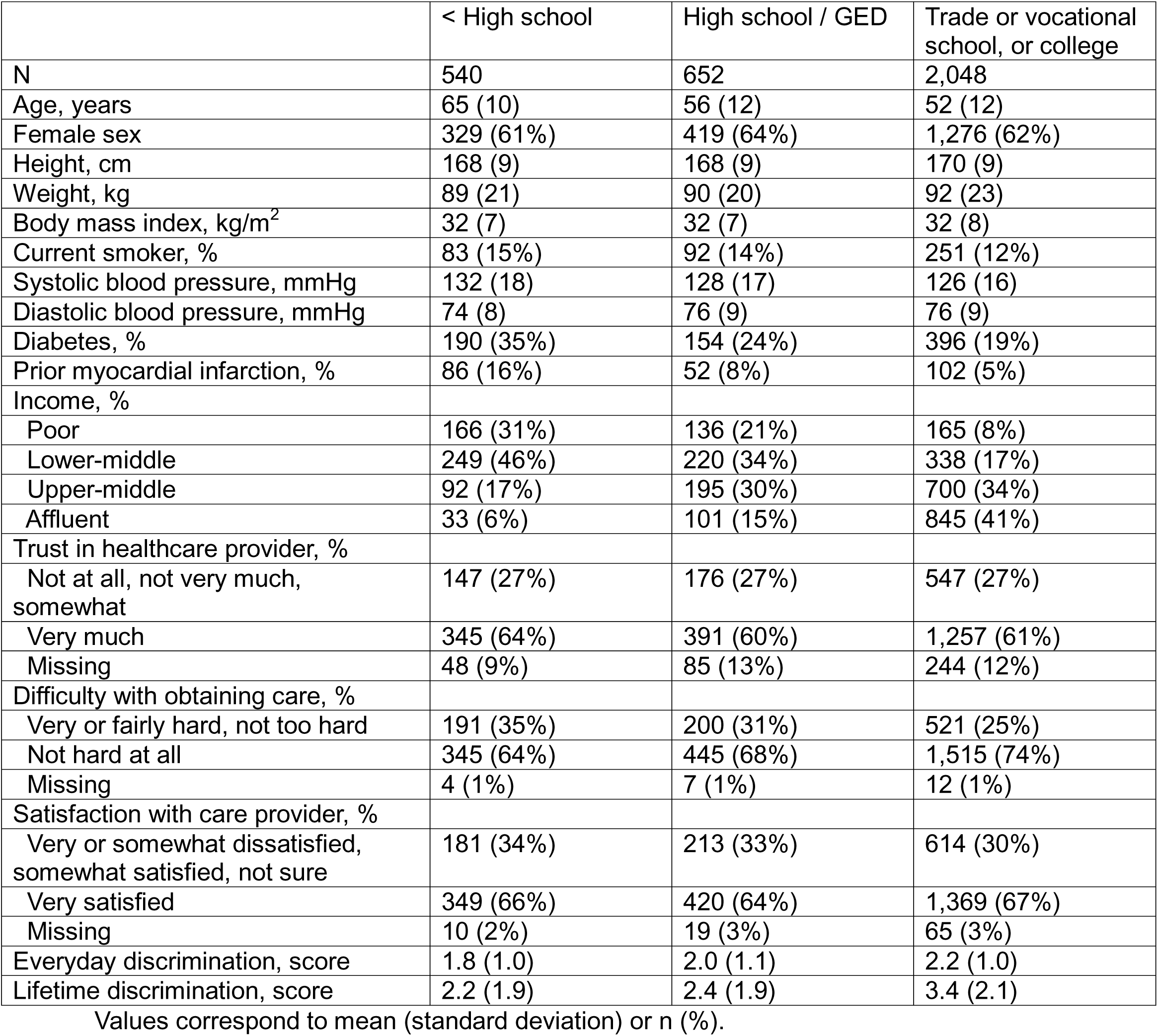
Baseline characteristics by education status, Jackson Heart Study, 2000-2004.

|  | < High school | High school / GED | Trade or vocational school, or college |
| --- | --- | --- | --- |
| N | 540 | 652 | 2,048 |
| Age, years | 65 (10) | 56 (12) | 52 (12) |
| Female sex | 329 (61%) | 419 (64%) | 1,276 (62%) |
| Height, cm | 168 (9) | 168 (9) | 170 (9) |
| Weight, kg | 89 (21) | 90 (20) | 92 (23) |
| Body mass index, kg/m<sup>2</sup> | 32 (7) | 32 (7) | 32 (8) |
| Current smoker, % | 83 (15%) | 92 (14%) | 251 (12%) |
| Systolic blood pressure, mmHg | 132 (18) | 128 (17) | 126 (16) |
| Diastolic blood pressure, mmHg | 74 (8) | 76 (9) | 76 (9) |
| Diabetes, % | 190 (35%) | 154 (24%) | 396 (19%) |
| Prior myocardial infarction, % | 86 (16%) | 52 (8%) | 102 (5%) |
| Income, % |  |  |  |
| Poor | 166 (31%) | 136 (21%) | 165 (8%) |
| Lower-middle | 249 (46%) | 220 (34%) | 338 (17%) |
| Upper-middle | 92 (17%) | 195 (30%) | 700 (34%) |
| Affluent | 33 (6%) | 101 (15%) | 845 (41%) |
| Trust in healthcare provider, % |  |  |  |
| Not at all, not very much, somewhat | 147 (27%) | 176 (27%) | 547 (27%) |
| Very much | 345 (64%) | 391 (60%) | 1,257 (61%) |
| Missing | 48 (9%) | 85 (13%) | 244 (12%) |
| Difficulty with obtaining care, % |  |  |  |
| Very or fairly hard, not too hard | 191 (35%) | 200 (31%) | 521 (25%) |
| Not hard at all | 345 (64%) | 445 (68%) | 1,515 (74%) |
| Missing | 4 (1%) | 7 (1%) | 12 (1%) |
| Satisfaction with care provider, % |  |  |  |
| Very or somewhat dissatisfied, somewhat satisfied, not sure | 181 (34%) | 213 (33%) | 614 (30%) |
| Very satisfied | 349 (66%) | 420 (64%) | 1,369 (67%) |
| Missing | 10 (2%) | 19 (3%) | 65 (3%) |
| Everyday discrimination, score | 1.8 (1.0) | 2.0 (1.1) | 2.2 (1.0) |
| Lifetime discrimination, score | 2.2 (1.9) | 2.4 (1.9) | 3.4 (2.1) |

Values correspond to mean (standard deviation) or n (%).

Higher educational attainment and income were associated with a lower risk of AF (**Tables 2 and 3**). In models adjusting for age, sex, income and insurance, having attended trade or vocational school, or college was associated with an 18% lower risk of AF compared to having less than a high school degree (HR 0.82, 95% CI 0.67, 1.02, Model 2). Associations were similar after further adjustment for healthcare access, experiences of discrimination, and clinical risk factors. A similar pattern was observed for income, with a 39% reduction in AF risk in the affluent group compared to individuals in the poor category (HR 0.61, 95% CI 0.47, 0.78). In analyses stratified by sex (female, male) or age (<55, ≥55 years of age), a stronger protective association between higher levels of education and AF risk was observed in younger participants (p for interaction = 0.01), with no differences by sex (**Figure 2**).

**Figure 2.**
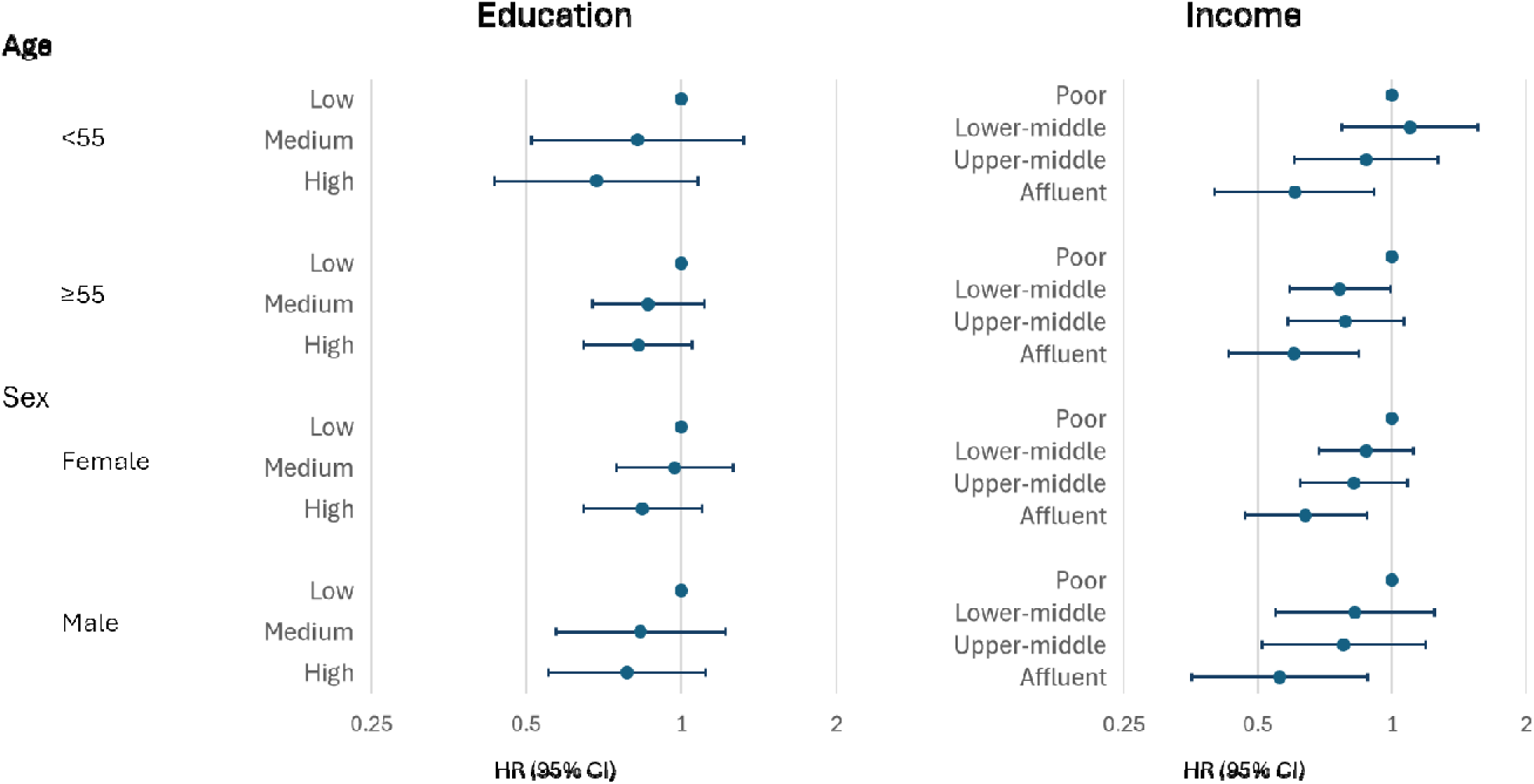
Association of education and income with incident atrial fibrillation stratified by age (<55, ≥55) and sex (female, male), Jackson Heart Study. Education was categorized as low (less than high school), medium (high school graduate/GED), and high (attended trade or vocational school, or college). Income was categorized by the study as poor, lower-middle, upper-middle, or affluent based on self-reported annual family income, family size, and poverty threshold for the year of the examination. Results from models adjusted by age and sex, as appropriate.

**Table 2.** Association of education with atrial fibrillation incidence, Jackson Heart Study, 2000-2016.

|  | < High school | High school / GED | Trade or vocational school, or college |  |
| --- | --- | --- | --- | --- |
| AF events, n | 184 | 185 | 448 |  |
| Person-years | 5,295 | 7,284 | 24,552 |  |
| Incidence rate* | 34.7 | 25.4 | 18.2 |  |
|  | HR (95% CI) |  |  | P for trend** |
| Model 1 | 1 (ref.) | 0.83 (0.67, 1.03) | 0.64 (0.53, 0.78) | <0.001 |
| Model 2 | 1 (ref.) | 0.92 (0.75, 1.15) | 0.82 (0.67, 1.02) | 0.06 |
| Model 3 | 1 (ref.) | 0.93 (0.75, 1.15) | 0.80 (0.65, 1.00) | 0.04 |
| Model 4 | 1 (ref.) | 0.96 (0.77, 1.19) | 0.84 (0.68, 1.04) | 0.08 |

\* Per 1,000 person-years. \*\* Categories modeled as ordinal variables. AF: atrial fibrillation. CI: confidence interval. HR: hazard ratio. Model 1: Cox regression model adjusted for age and sex. Model 2: Model 1 + additional adjustment for income and type of insurance. Model 3: Model 2 + additional adjustment for everyday discrimination, lifetime discrimination, healthcare access variables (trust in provider, difficulty accessing care, satisfaction with care). Model 4: Model 3 + additional adjustment for smoking, body mass index, height, use of blood pressure medications, systolic blood pressure, diastolic blood pressure, diabetes, and prevalent myocardial infarction.

**Table 3.**
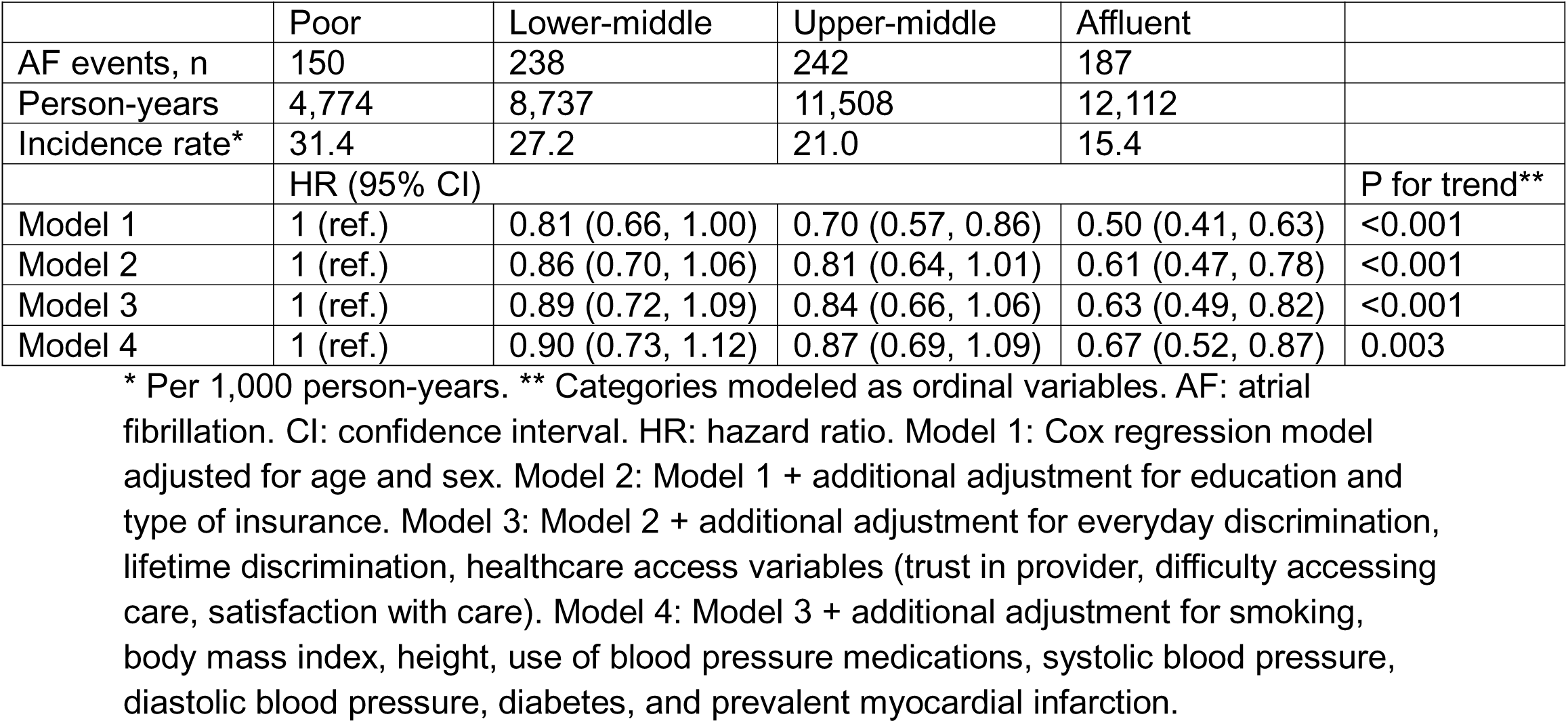
Association of income with atrial fibrillation incidence, Jackson Heart Study, 2000-2016.

Among measures of healthcare access, public insurance was associated with a higher risk of AF compared to being uninsured (HR 1.27, 95% CI 0.99, 1.63, Model 2), with no associations of private or dual public-private insurance with AF risk (**Table 4**). Better self-reported access to healthcare and greater satisfaction with care provider was associated with lower AF risk (**Table 5**). Individuals reporting no difficulty obtaining care (‘not hard at all’) had a 16% lower risk of AF than those in other categories (HR 0.84, 95% CI 0.72, 0.97, Model 2), while being very satisfied with the care provider was associated with a 10% lower AF risk (HR 0.90, 95% CI 0.77, 1.04). Trust in healthcare provider was not associated with AF risk.

**Table 4.** Association of insurance type with atrial fibrillation incidence, Jackson Heart Study, 2000-2016.

|  | Uninsured | Private | Public | Dual private-public |
| --- | --- | --- | --- | --- |
| AF events, n | 109 | 357 | 225 | 126 |
| Person-years | 5,038 | 20,293 | 6,333 | 5,468 |
| Incidence rate* | 21.6 | 17.6 | 35.5 | 23.0 |
|  | HR (95% CI) |  |  |  |
| Model 1 | 1 (ref.) | 0.82 (0.66, 1.02) | 1.31 (1.02, 1.69) | 0.86 (0.65, 1.13) |
| Model 2 | 1 (ref.) | 1.00 (0.80, 1.26) | 1.27 (0.99, 1.63) | 1.02 (0.77, 1.35) |
| Model 3 | 1 (ref.) | 1.01 (0.80, 1.28) | 1.27 (0.99, 1.63) | 1.03 (0.77, 1.37) |
| Model 4 | 1 (ref.) | 1.04 (0.82, 1.31) | 1.24 (0.96, 1.59) | 1.04 (0.78, 1.39) |

\* Per 1,000 person-years. AF: atrial fibrillation. CI: confidence interval. HR: hazard ratio. Model 1: Cox regression model adjusted for age and sex. Model 2: Model 1 + additional adjustment for education and income. Model 3: Model 2 + additional adjustment for everyday discrimination, lifetime discrimination, healthcare access variables (trust in provider, difficulty accessing care, satisfaction with care). Model 4: Model 3 + additional adjustment for smoking, body mass index, height, use of blood pressure medications, systolic blood pressure, diastolic blood pressure, diabetes, and prevalent myocardial infarction.

**Table 5.** Association of healthcare access-related variables with atrial fibrillation incidence, Jackson Heart Study, 2000-2016.

|  | Trust in healthcare provider |  | Difficulty with obtaining care |  | Satisfaction with care provider |  |
| --- | --- | --- | --- | --- | --- | --- |
|  | Not at all, not very much, somewhat | Very much | Very or fairly hard, not too hard | Not hard at all | Very or somewhat dissatisfied, somewhat satisfied, not sure | Very satisfied |
| AF events, n | 233 | 513 | 271 | 542 | 276 | 527 |
| Person-years | 10,024 | 22,607 | 10,116 | 26,726 | 11,505 | 24,419 |
| Incidence rate* | 23.2 | 22.7 | 26.8 | 20.3 | 24.0 | 21.6 |
|  | HR (95% CI) |  |  |  |  |  |
| Model 1 | 1 (ref.) | 0.92 (0.79, 1.07) | 1 (ref.) | 0.76 (0.65, 0.87) | 1 (ref.) | 0.84 (0.73, 0.98) |
| Model 2 | 1 (ref.) | 0.95 (0.81, 1.11) | 1 (ref.) | 0.84 (0.72, 0.97) | 1 (ref.) | 0.90 (0.77, 1.04) |
| Model 3 | 1 (ref.) | 1.04 (0.85, 1.26) | 1 (ref.) | 0.88 (0.75, 1.03) | 1 (ref.) | 0.93 (0.77, 1.12) |
| Model 4 | 1 (ref.) | 1.00 (0.82, 1.22) | 1 (ref.) | 0.90 (0.76, 1.05) | 1 (ref.) | 0.94 (0.77, 1.13) |

\* Per 1,000 person-years. AF: atrial fibrillation. CI: confidence interval. HR: hazard ratio. Model 1: Cox regression model adjusted for age and sex. Model 2: Model 1 + additional adjustment for education, income, and type of insurance. Model 3: Model 2 + additional adjustment for everyday discrimination, lifetime discrimination, healthcare access variables (trust in provider, difficulty accessing care, satisfaction with care), excluding the variable evaluated as primary exposure. Model 4: Model 3 + additional adjustment for smoking, body mass index, height, use of blood pressure medications, systolic blood pressure, diastolic blood pressure, diabetes, and prevalent myocardial infarction.

Overall findings were similar when we used a more specific AF definition requiring a self-reported AF diagnosis in at least 2 follow-up surveys (**Supplemental Tables S2-S5**) or when answers to healthcare access-related questions were disaggregated in more categories (**Supplemental Tables S6-S8**).

## DISCUSSION

This analysis of a large cohort of Black Americans found that both higher socioeconomic position and better healthcare access were associated with lower risk of AF. Results were generally consistent across multiple indicators of socioeconomic position, including educational attainment and income. By focusing exclusively on Black Americans, this study evaluated socioeconomic gradients in AF risk without relying on comparisons across racial groups, which can be difficult to interpret because race reflects a complex constellation of social, economic, environmental, and historical experiences, intimately intertwined with individual socioeconomic position.^17^ As a result, our findings clarify the role of socioeconomic factors in AF risk within a population that has been underrepresented in prior epidemiologic studies.

Numerous previous studies have reported a lower risk of AF among individuals with higher socioeconomic status or those living in less deprived neighborhoods.^8, 9, 18–21^ Our findings are consistent with this literature while minimizing race-related confounding by focusing exclusively on Black individuals.

Healthcare access also emerged as a potential contributor to AF risk. Prior evidence regarding the influence of healthcare access and quality on AF risk is extremely limited. Greater trust in and satisfaction with healthcare providers have been associated with higher adherence to oral anticoagulation and better quality of life among patients with AF,^22, 23^ and AF catheter ablation rates are lower in individuals with public insurance or no insurance than among those with private insurance.^24^ However, no prior studies have reported AF risk according to insurance status or other measures of healthcare access.

Multiple pathways likely link socioeconomic status to AF risk,^25^ including a higher burden of cardiometabolic disease, adverse health behaviors, chronic psychosocial stress, environmental exposures, and reduced access to high-quality preventive healthcare among individuals with lower socioeconomic position.^26^ At the same time, disparities in healthcare access and utilization due to education, income or insurance status may influence the likelihood that AF is detected, potentially resulting in more diagnoses among individuals with better healthcare access.^5^ Our findings suggest that, within this cohort of Black adults, socioeconomic disadvantage and poorer healthcare access were not associated with lower observed AF incidence, as might be expected if underascertainment related to these factors predominated. The described socioeconomic pattern is comparable to that observed for other cardiovascular diseases.^27^

These findings also offer additional insight into the so-called “racial paradox” in AF.^28^ Extensive epidemiologic evidence has consistently shown that Black individuals have a lower risk of AF than White individuals despite a greater burden of established AF risk factors.^2, 29^ Prior reviews have suggested that disparities in access to high-quality healthcare among Black individuals could contribute to this phenomenon.^5, 26, 30^ Our findings, showing that higher socioeconomic status and better healthcare access were associated with lower rather than higher observed AF risk within Black adults, argue against a simple explanation in which socioeconomic disadvantage alone produces lower AF rates through underascertainment. However, because our analysis did not compare Black and White individuals, it cannot directly assess differential AF ascertainment between racial groups. Alternative potential explanations include differences in genetic ancestry, with a higher risk of AF among individuals with a greater proportion of European genetic ancestry,^6, 7^ as well as selective survival, whereby higher mortality rates among Black versus White individuals selectively reduce the number of people who survive long enough to develop AF.

Strengths of this study include the inclusion of a community-based sample, the geographic and racial homogeneity, which reduces the likelihood of confounding, a relatively large sample size with an adequate number of AF events, and the availability of multiple measures of socioeconomic status and healthcare access. Limitations include the ascertainment of AF, which relied primarily on self-report, resulting in outcome misclassification, the exclusion of participants who did not consent to data sharing with outside investigators, which could introduce selection bias, and the inability to generalize these findings to other populations.

Future studies should move beyond documenting socioeconomic disparities in AF to identifying the mechanisms underlying these associations so that appropriate preventive strategies can be developed and implemented. Longitudinal investigations adopting a life-course perspective and integrating repeated measures of socioeconomic position, psychosocial stress, cardiometabolic risk factors, biomarkers, and environmental exposures may clarify the biological pathways through which socioeconomic disadvantage contributes to AF. Studies incorporating systematic rhythm monitoring may also help distinguish true differences in AF incidence from differential detection across socioeconomic groups. Finally, additional research is needed to elucidate the causes explaining the racial paradox in AF epidemiology.

In conclusion, higher socioeconomic position and better healthcare access were associated with a lower risk of AF in a cohort of Black adults from the American South. These findings underscore the importance of understanding the mechanisms linking socioeconomic disadvantage and healthcare access with AF and may help identify opportunities for future preventive interventions. The results also argue against a simple explanation in which socioeconomic disadvantage and poorer healthcare access result in lower observed AF incidence through underascertainment within Black adults. However, they do not exclude differential ascertainment between Black and White populations as a contributor to racial differences in AF incidence.

## Supporting information

Supplemental results

## Data Availability

All data used in the present study are available online at NHLBI BioLINCC

https://biolincc.nhlbi.nih.gov/home/

## ACKNOWLEDGEMENTS

This manuscript was prepared using JHS Research Materials obtained from the National Heart, Lung, and Blood Institute (NHLBI) Biologic Specimen and Data Repository Information Coordinating Center (BioLINCC) and does not necessarily reflect the opinions or views of the JHS or the NHLBI.

