## Supplemental results for "Socioeconomic status, healthcare access, and risk of atrial fibrillation: an investigation using the Jackson Heart Study Research Materials"

Supplemental Table S1. Baseline characteristics by income.

|  | Poor | Lower-middle | Upper-middle | Affluent |
| --- | --- | --- | --- | --- |
| N | 467 | 807 | 987 | 979 |
| Age, years | 56 (15) | 58 (13) | 53 (12) | 54 (11) |
| Female sex | 350 (75%) | 546 (68%) | 618 (63%) | 510 (52%) |
| Height, cm | 167 (9) | 168 (9) | 169 (9) | 171 (10) |
| Weight, kg | 91 (24) | 91 (22) | 92 (22) | 91 (20) |
| Body mass index, kg/m^2^ | 33 (9) | 32 (8) | 32 (7) | 31 (6) |
| Current smoker, % | 81 (17%) | 126 (16%) | 136 (14%) | 83 (8.5%) |
| Systolic blood pressure, mmHg | 129 (17) | 129 (18) | 127 (16) | 126 (15) |
| Diastolic blood pressure, mmHg | 75 (9) | 75 (9) | 76 (9) | 76 (8) |
| Diabetes, % | 131 (28%) | 214 (27%) | 229 (23%) | 166 (17%) |
| Prior myocardial infarction, % | 58 (12%) | 75 (9.3%) | 59 (6.0%) | 48 (4.9%) |
| Education, % |  |  |  |  |
| < High school | 166 (36%) | 249 (31%) | 92 (9.3%) | 33 (3.4%) |
| High school / GED | 136 (29%) | 220 (27%) | 195 (20%) | 101 (10%) |
| Trade or vocational school, or college | 165 (35%) | 338 (42%) | 700 (71%) | 845 (86%) |
| Trust in healthcare provider, % |  |  |  |  |
| Not at all, not very much, somewhat | 146 (31%) | 212 (26%) | 271 (27%) | 241 (25%) |
| Very much | 262 (56%) | 494 (61%) | 591 (60%) | 646 (66%) |
| Missing | 59 (13%) | 101 (13%) | 125 (13%) | 92 (9%) |
| Difficulty with obtaining care, % |  |  |  |  |
| Very or fairly hard, not too hard | 226 (48%) | 281 (35%) | 242 (25%) | 163 (17%) |
| Not hard at all | 239 (51%) | 518 (64%) | 743 (75%) | 805 (82%) |
| Missing | 2 (<1%) | 8 (1%) | 2 (<1%) | 11 (1%) |
| Satisfaction with care provider, % |  |  |  |  |
| Very or somewhat dissatisfied, somewhat satisfied, not sure | 193 (41%) | 243 (30%) | 315 (32%) | 257 (26%) |
| Very satisfied | 261 (56%) | 539 (67%) | 641 (65%) | 697 (71%) |
| Missing | 13 (3%) | 25 (3%) | 31 (3%) | 25 (3%) |
| Everyday discrimination, score | 2.1 (1.1) | 2.1 (1.1) | 2.1 (1.0) | 2.1 (0.9) |
| Lifetime discrimination, score | 2.7 (2.0) | 2.6 (2.0) | 3.1 (2.1) | 3.3 (2.1) |

Values correspond to mean (standard deviation) or n (%).

Supplemental Table S2. Association of education with atrial fibrillation incidence, Jackson Heart Study, 2000-2016. Analysis using AF definition requiring at least two self-reports at different times.

|  | < High school | High school / GED | Trade or vocational school, or college |  |
| --- | --- | --- | --- | --- |
| AF events, n | 95 | 100 | 233 |  |
| Person-years | 6,016 | 8,040 | 26,396 |  |
| Incidence rate* | 15.8 | 12.4 | 8.8 |  |
|  | HR (95% CI) | | | P for trend |
| Model 1 | 1 (ref.) | 0.83 (0.62, 1.11) | 0.61 (0.47, 0.79) | <0.001 |
| Model 2 | 1 (ref.) | 0.91 (0.68, 1.22) | 0.76 (0.57, 1.02) | 0.05 |
| Model 3 | 1 (ref.) | 0.93 (0.69, 1.25) | 0.74 (0.55, 1.00) | 0.03 |
| Model 4 | 1 (ref.) | 0.96 (0.71, 1.30) | 0.78 (0.58, 1.06) | 0.07 |

* Per 1,000 person-years. AF: atrial fibrillation. CI: confidence interval. HR: hazard ratio. Model 1: Cox regression model adjusted for age and sex. Model 2: Model 1 + additional adjustment for income and type of insurance. Model 3: Model 2 + additional adjustment for everyday discrimination, lifetime discrimination, healthcare access variables (trust in provider, difficulty accessing care, satisfaction with care). Model 4: Model 3 + additional adjustment for smoking, body mass index, height, use of blood pressure medications, systolic blood pressure, diastolic blood pressure, diabetes, and prevalent myocardial infarction.

Supplemental Table S3. Association of income with atrial fibrillation incidence, Jackson Heart Study, 2000-2016. Analysis using AF definition requiring at least two self-reports at different times.

|  | Poor | Lower-middle | Upper-middle | Affluent |  |
| --- | --- | --- | --- | --- | --- |
| AF events, n | 78 | 128 | 123 | 99 |  |
| Person-years | 5,372 | 9,674 | 12,507 | 12,900 |  |
| Incidence rate* | 14.5 | 13.2 | 9.8 | 7.7 |  |
|  | HR (95% CI) | | | | P for trend |
| Model 1 | 1 (ref.) | 0.88 (0.67, 1.17) | 0.70 (0.53, 0.94) | 0.55 (0.41, 0.74) | <0.001 |
| Model 2 | 1 (ref.) | 0.94 (0.70, 1.25) | 0.83 (0.60, 1.14) | 0.68 (0.48, 0.97) | 0.02 |
| Model 3 | 1 (ref.) | 0.98 (0.74, 1.31) | 0.89 (0.64, 1.22) | 0.73 (0.51, 1.05) | 0.06 |
| Model 4 | 1 (ref.) | 1.01 (0.76, 1.35) | 0.92 (0.66, 1.27) | 0.79 (0.55, 1.13) | 0.14 |

* Per 1,000 person-years. AF: atrial fibrillation. CI: confidence interval. HR: hazard ratio. Model 1: Cox regression model adjusted for age and sex. Model 2: Model 1 + additional adjustment for education and type of insurance. Model 3: Model 2 + additional adjustment for everyday discrimination, lifetime discrimination, healthcare access variables (trust in provider, difficulty accessing care, satisfaction with care). Model 4: Model 3 + additional adjustment for smoking, body mass index, height, use of blood pressure medications, systolic blood pressure, diastolic blood pressure, diabetes, and prevalent myocardial infarction.

Supplemental Table S4. Association of insurance type with atrial fibrillation incidence, Jackson Heart Study, 2000-2016. Analysis using AF definition requiring at least two self-reports at different times.

|  | Uninsured | Private | Public | Dual private-public |
| --- | --- | --- | --- | --- |
| AF events, n | 61 | 188 | 111 | 68 |
| Person-years | 5,457 | 21,800 | 7,206 | 5,989 |
| Incidence rate* | 11.2 | 8.6 | 15.4 | 11.4 |
|  | HR (95% CI) | | | |
| Model 1 | 1 (ref.) | 0.78 (0.58, 1.04) | 1.26 (0.89, 1.78) | 0.94 (0.65, 1.36) |
| Model 2 | 1 (ref.) | 0.94 (0.69, 1.29) | 1.22 (0.86, 1.72) | 1.11 (0.76, 1.62) |
| Model 3 | 1 (ref.) | 0.98 (0.71, 1.33) | 1.22 (0.87, 1.73) | 1.13 (0.77, 1.67) |
| Model 4 | 1 (ref.) | 1.00 (0.73, 1.38) | 1.17 (0.83, 1.65) | 1.16 (0.79, 1.71) |

* Per 1,000 person-years. AF: atrial fibrillation. CI: confidence interval. HR: hazard ratio. Model 1: Cox regression model adjusted for age and sex. Model 2: Model 1 + additional adjustment for education and income. Model 3: Model 2 + additional adjustment for everyday discrimination, lifetime discrimination, healthcare access variables (trust in provider, difficulty accessing care, satisfaction with care). Model 4: Model 3 + additional adjustment for smoking, body mass index, height, use of blood pressure medications, systolic blood pressure, diastolic blood pressure, diabetes, and prevalent myocardial infarction.

Supplemental Table S5. Association of healthcare access-related variables with atrial fibrillation incidence, Jackson Heart Study, 2000-2016. Analysis using AF definition requiring at least two self-reports at different times.

|  | Trust in healthcare provider | | Difficulty with obtaining care | | | Satisfaction with care provider | |
| --- | --- | --- | --- | --- | --- | --- | --- |
|  | Not at all, not very much, somewhat | Very much | Very or fairly hard, not too hard | Not hard at all | Very or somewhat dissatisfied, somewhat satisfied | | Very satisfied |
| AF events, n | 128 | 269 | 156 | 271 | 151 | | 273 |
| Person-years | 10,964 | 24,636 | 11,120 | 29,021 | 12,612 | | 26,560 |
| Incidence rate* | 11.7 | 10.9 | 14.0 | 9.3 | 12.0 | | 10.3 |
|  | HR (95% CI) | | | | | | |
| Model 1 | 1 (ref.) | 0.90 (0.73, 1.11) | 1 (ref.) | 0.66 (0.54, 0.81) | 1 (ref.) | | 0.81 (0.67, 0.99) |
| Model 2 | 1 (ref.) | 0.93 (0.75, 1.15) | 1 (ref.) | 0.72 (0.59, 0.89) | 1 (ref.) | | 0.85 (0.70, 1.05) |
| Model 3 | 1 (ref.) | 1.05 (0.80, 1.37) | 1 (ref.) | 0.76 (0.62, 0.95) | 1 (ref.) | | 0.91 (0.70, 1.18) |
| Model 4 | 1 (ref.) | 1.01 (0.77, 1.32) | 1 (ref.) | 0.78 (0.63, 0.98) | 1 (ref.) | | 0.92 (0.71, 1.20) |

* Per 1,000 person-years. AF: atrial fibrillation. CI: confidence interval. HR: hazard ratio. Model 1: Cox regression model adjusted for age and sex. Model 2: Model 1 + additional adjustment for education, income, and type of insurance. Model 3: Model 2 + additional adjustment for everyday discrimination, lifetime discrimination, healthcare access variables (trust in provider, difficulty accessing care, satisfaction with care), excluding the variable evaluated as primary exposure. Model 4: Model 3 + additional adjustment for smoking, body mass index, height, use of blood pressure medications, systolic blood pressure, diastolic blood pressure, diabetes, and prevalent myocardial infarction.

Supplemental Tables S6. Association of trust in healthcare providers with atrial fibrillation incidence, Jackson Heart Study, 2000-2016.

|  | How much do you trust your usual healthcare provider? | | |  |
| --- | --- | --- | --- | --- |
|  | Not at all / not very much** | Somewhat | Very much |  |
| AF events, n | 39 | 194 | 513 |  |
| Person-years | 1,140 | 8,884 | 22,607 |  |
| Incidence rate* | 34.2 | 21.8 | 22.7 |  |
|  | HR (95%CI) |  |  | P for trend*** |
| Model 1 | 1 (ref.) | 0.65 (0.46, 0.92) | 0.64 (0.46, 0.89) | 0.07 |
| Model 2 | 1 (ref.) | 0.70 (0.49, 0.98) | 0.70 (0.50, 0.97) | 0.23 |
| Model 3 | 1 (ref.) | 0.82 (0.54, 1.25) | 0.91 (0.59, 1.40) | 0.78 |
| Model 4 | 1 (ref.) | 0.85 (0.56, 1.30) | 0.91 (0.59, 1.40) | 0.97 |

* Per 1,000 person-years. ** Combined due to small number of events in “not at all” category. *** Categories modeled as ordinal variables. AF: atrial fibrillation. CI: confidence interval. HR: hazard ratio. Model 1: Cox regression model adjusted for age and sex. Model 2: Model 1 + additional adjustment for education, income, and type of insurance. Model 3: Model 2 + additional adjustment for everyday discrimination, lifetime discrimination, other healthcare access variables (difficulty accessing care, satisfaction with care). Model 4: Model 3 + additional adjustment for smoking, body mass index, height, use of blood pressure medications, systolic blood pressure, diastolic blood pressure, diabetes, and prevalent myocardial infarction.

Supplemental Table S7. Association of difficulty in accessing healthcare with atrial fibrillation incidence, Jackson Heart Study, 2000-2016.

|  | How hard has it been for you to get health services you have needed? | | | |  |
| --- | --- | --- | --- | --- | --- |
|  | Very hard | Fairly hard | Not too hard | Not hard at all |  |
| AF events, n | 65 | 63 | 143 | 542 |  |
| Person-years | 1,827 | 2,012 | 6,277 | 26,726 |  |
| Incidence rate* | 35.6 | 31.3 | 22.8 | 20.3 |  |
|  | HR (95% CI) | | | | P for trend** |
| Model 1 | 1 (ref.) | 0.88 (0.62, 1.25) | 0.62 (0.47, 0.84) | 0.56 (0.43, 0.72) | <0.001 |
| Model 2 | 1 (ref.) | 0.93 (0.65, 1.32) | 0.66 (0.49, 0.89) | 0.64 (0.49, 0.84) | <0.001 |
| Model 3 | 1 (ref.) | 0.88 (0.61, 1.28) | 0.62 (0.46, 0.86) | 0.62 (0.46, 0.83) | <0.001 |
| Model 4 | 1 (ref.) | 0.87 (0.60, 1.26) | 0.64 (0.46, 0.87) | 0.63 (0.47, 0.85) | 0.002 |

* Per 1,000 person-years. ** Categories modeled as ordinal variables. AF: atrial fibrillation. CI: confidence interval. HR: hazard ratio. Model 1: Cox regression model adjusted for age and sex. Model 2: Model 1 + additional adjustment for education, income, and type of insurance. Model 3: Model 2 + additional adjustment for everyday discrimination, lifetime discrimination, other healthcare access variables (trust in provider, satisfaction with care). Model 4: Model 3 + additional adjustment for smoking, body mass index, height, use of blood pressure medications, systolic blood pressure, diastolic blood pressure, diabetes, and prevalent myocardial infarction.

Supplemental Table S8. Association of satisfaction with healthcare provider with atrial fibrillation incidence, Jackson Heart Study, 2000-2016

|  | How satisfied are you with your regular (or most recent) doctor or health professional?* | | |  |
| --- | --- | --- | --- | --- |
|  | Very / somewhat dissatisfied*** | Somewhat satisfied | Very satisfied |  |
| AF events, n | 36 | 228 | 527 |  |
| Person-years | 1,264 | 9,520 | 24,419 |  |
| Incidence rate** | 28.5 | 23.9 | 21.6 |  |
|  | HR (95% CI) | | | P for trend**** |
| Model 1 | 1 (ref.) | 0.77 (0.54, 1.09) | 0.65 (0.47, 0.92) | 0.004 |
| Model 2 | 1 (ref.) | 0.80 (0.56, 1.14) | 0.72 (0.51, 1.01) | 0.04 |
| Model 3 | 1 (ref.) | 0.97 (0.62, 1.52) | 0.92 (0.57, 1.47) | 0.68 |
| Model 4 | 1 (ref.) | 0.93 (0.60, 1.46) | 0.90 (0.56, 1.44) | 0.72 |

* 62 participants answering ‘Not sure’ to this question were excluded from this analysis. ** Per 1,000 person-years. *** Combined due to small number of events in these categories. **** Categories modeled as ordinal variables. AF: atrial fibrillation. CI: confidence interval. HR: hazard ratio. Model 1: Cox regression model adjusted for age and sex. Model 2: Model 1 + additional adjustment for education, income, and type of insurance. Model 3: Model 2 + additional adjustment for everyday discrimination, lifetime discrimination, other healthcare access variables (trust in healthcare provider, difficulty accessing care). Model 4: Model 3 + additional adjustment for smoking, body mass index, height, use of blood pressure medications, systolic blood pressure, diastolic blood pressure, diabetes, and prevalent myocardial infarction.
